# Perceptions and Experiences of Healthcare Professionals Regarding Green Practices to Promote Environmental Sustainability in Health Services: A Protocol for a Systematic Review of Qualitative Evidence

**DOI:** 10.1101/2025.07.06.25330984

**Authors:** Ravi Shankar, Fiona Devi, Xu Qian

## Abstract

**Background:** The healthcare sector significantly contributes to environmental degradation and climate change. Implementing sustainable practices, known as “green practices,” can mitigate these negative impacts. Healthcare professionals play a crucial role in adopting and promoting green practices, but their perceptions and experiences are not well understood.

**Objective:** This systematic review protocol outlines methods for synthesizing qualitative evidence on healthcare professionals’ perceptions and experiences regarding green practices in health services. The review aims to identify facilitators, barriers, and strategies for implementing green practices from healthcare professionals’ perspectives.

**Methods:** We will search PubMed, Web of Science, Embase, CINAHL, MEDLINE, The Cochrane Library, PsycINFO, and Scopus from each database’s inception to July 2025. We will include qualitative studies exploring healthcare professionals’ perceptions and experiences regarding green practices in health services. Two reviewers will independently screen studies using Covidence, extract data, and assess methodological quality using the Critical Appraisal Skills Programme (CASP) checklist. We will use thematic synthesis to analyze findings. Risk of bias will be assessed using the Joanna Briggs Institute (JBI) Critical Appraisal Checklist for Qualitative Research.

**Discussion:** This review will provide insights into healthcare professionals’ perceptions and experiences regarding green practices. Findings will inform strategies for implementing sustainable practices in healthcare, considering key stakeholders’ perspectives. This may contribute to reducing healthcare’s environmental impact and improving planetary health. Limitations include English language restriction and potential non-generalizability of qualitative evidence.

## Introduction

### Background

The healthcare sector is a significant contributor to environmental degradation and climate change. In the United States, healthcare accounts for nearly 10% of the country’s greenhouse gas emissions and 9% of harmful non-greenhouse air pollutants [1]. On a global scale, if the health sector were a country, it would be the fifth-largest emitter of greenhouse gases [2]. Healthcare facilities also generate substantial amounts of waste, with the U.S. healthcare system alone producing an estimated 4.67 million tons of medical waste annually [3].

To mitigate these negative environmental impacts, there is growing recognition of the need to implement sustainable practices in healthcare, often referred to as “green practices” [4]. Green practices encompass a wide range of strategies aimed at reducing the ecological footprint of healthcare activities while maintaining or improving the quality of care delivered to patients [5]. Examples of green practices include reducing energy and water consumption, minimizing waste generation, promoting recycling and reuse, using environmentally friendly products, and encouraging active transport among staff and patients [6].

Adopting green practices in healthcare settings not only benefits the environment but can also lead to cost savings, improved patient outcomes, and better population health [7]. As the impacts of climate change on human health become increasingly apparent, healthcare professionals have a responsibility to lead by example in promoting sustainability and mitigating the sector’s environmental impact [8]. The Hippocratic Oath, which underpins the ethical principles of the medical profession, has been interpreted by some to encompass a duty to protect the environment as a means of safeguarding human health [9].

Healthcare professionals, including doctors, nurses, pharmacists, and healthcare managers, play a vital role in implementing green practices within their organizations [10]. Their knowledge, attitudes, and behaviors regarding sustainability can significantly influence the adoption and success of green initiatives in healthcare settings [11]. Therefore, understanding healthcare professionals’ perceptions and experiences is crucial for designing effective interventions and policies to promote sustainable practices in health services.

Several studies have explored healthcare professionals’ perspectives on environmental sustainability in healthcare settings [12-14]. A survey of nurses in the United Kingdom found that while most agreed that sustainability is important for health services, they perceived significant barriers such as lack of time and inadequate recycling facilities [12]. Interviews with hospital managers in Italy revealed that although they recognized the importance of sustainability, competing priorities and limited resources often hindered the implementation of green practices [13]. A focus group study with anesthesiologists in Australia identified strategies for reducing waste and energy consumption in the operating room, such as using low-flow anesthetic techniques and turning off unused equipment when not in use [14].

However, the existing evidence on healthcare professionals’ perceptions and experiences regarding green practices is fragmented and has not been synthesized in a systematic manner. A comprehensive systematic review is needed to critically appraise and integrate the qualitative findings from diverse healthcare contexts. Understanding the facilitators, barriers, and strategies for implementing green practices from the perspective of healthcare professionals is essential for developing targeted interventions and policies to promote sustainability in health services.

### Objective

The objective of this systematic review is to synthesize qualitative evidence on healthcare professionals’ perceptions and experiences regarding green practices in health services. Specifically, we aim to:

1. Identify the perceived facilitators and barriers to implementing green practices in healthcare settings from the perspective of healthcare professionals.
2. Explore healthcare professionals’ experiences with adopting and promoting green practices in their work environments.
3. Synthesize strategies and recommendations for implementing green practices in health services based on insights from healthcare professionals.

### Methods

This systematic review protocol follows the Preferred Reporting Items for Systematic Review and Meta-Analysis Protocols (PRISMA-P) guidelines [15]. The completed review will adhere to the Enhancing Transparency in Reporting the Synthesis of Qualitative Research (ENTREQ) statement [16].

### Review question (PICO)

Using the PICO (Population, Interest, Context, Outcome) framework, the review question is:

What are the perceptions and experiences of healthcare professionals (P) regarding green practices (I) in health services (Co) and what are the perceived facilitators, barriers, and strategies for implementing these practices (O)?

### Eligibility criteria

We will select studies for inclusion in this review according to the following criteria:

### Types of studies

We will include primary qualitative studies that use recognized methods for qualitative data collection and analysis, such as interviews, focus groups, ethnography, or qualitative surveys. We will exclude studies that collected data using qualitative methods but did not analyze the data qualitatively (e.g., open-ended survey responses analyzed using quantitative methods). Mixed-methods studies will be included if they report qualitative findings separately. We will exclude commentaries, editorials, reviews, and other non-primary research articles.

### Types of participants

We will include studies that focus on healthcare professionals, defined as individuals who provide clinical care or support the delivery of health services. This includes but is not limited to doctors, nurses, pharmacists, allied health professionals, healthcare managers, and support staff working in healthcare settings. We will exclude studies that only focus on students or trainees who are not yet qualified healthcare professionals.

### Types of settings

We will include studies conducted in any healthcare setting, including hospitals, clinics, primary care centers, community health services, pharmacies, and aged care facilities. We will exclude studies conducted exclusively in educational institutions or research centers that do not involve the delivery of health services.

### Types of green practices

We will include studies that explore healthcare professionals’ perceptions and experiences regarding green practices, defined as strategies, interventions, or initiatives aimed at promoting environmental sustainability in health services. Examples of green practices include but are not limited to:

- Reducing energy consumption through energy-efficient technologies, renewable energy sources, or behavior change interventions
- Conserving water through efficient fixtures, reuse of greywater, or rainwater harvesting
- Minimizing waste generation by reducing packaging, implementing reusable medical devices, or segregating waste streams
- Promoting recycling and reuse of materials such as paper, plastic, glass, and metal
- Using environmentally friendly products such as non-toxic cleaning agents, biodegradable disposables, or products with reduced packaging
- Encouraging active transport (e.g., walking, cycling) or the use of public transport among staff and patients to reduce carbon emissions from travel
- Serving sustainably sourced, locally produced, or plant-based food options in healthcare facilities
- Implementing telehealth or virtual care services to reduce the need for patient travel

We will exclude studies that focus solely on sustainability practices in healthcare education or research settings, without considering the application of these practices to clinical care delivery or health service operations.

### Time frame

We will include studies published from each database’s inception until the date of our final search, which is planned for July 2025. This broad time frame is chosen to capture all relevant studies on the topic, considering that the concept of green practices in healthcare has evolved over time.

### Language

We will include only studies published in English, due to resource constraints and the language capabilities of the review team. We acknowledge this as a limitation of the review, as we may potentially exclude relevant studies published in other languages.

### Information sources and search strategy

We will conduct a comprehensive search of the following electronic bibliographic databases: PubMed, Web of Science, Embase, CINAHL (Cumulative Index to Nursing and Allied Health Literature), MEDLINE (via Ovid), the Cochrane Library, PsycINFO (via Ovid), and Scopus.

The search strategy will be developed in consultation with a health sciences librarian and will combine terms related to three key concepts: (1) healthcare professionals, (2) perceptions and experiences regarding green practices, and (3) qualitative research. We will use a combination of free-text keywords and controlled vocabulary terms (e.g., Medical Subject Headings [MeSH] in PubMed) to capture relevant studies. The search strategy will be adapted for each database, taking into account differences in indexing and syntax.

A draft search string template is provided below:

((“health personnel” OR “healthcare professional*” OR “health care professional*” OR “healthcare worker*” OR “health care worker*” OR “medical staff” OR “allied health” OR doctor* OR physician* OR nurs* OR pharmacist*) AND (“green practice*” OR “sustainable practice*” OR “environmentally sustainable” OR “environmentally friendly” OR “eco-friendly” OR “ecological footprint” OR “carbon footprint” OR “environmental impact” OR “environmental stewardship” OR “greening healthcare” OR “green hospital*” OR “sustainable healthcare” OR “sustainable health care” OR recycl* OR “energy conservation” OR “renewable energy” OR “waste management” OR “active transport” OR “public transport” OR telehealth) AND (perception* OR perspective* OR view* OR opinion* OR attitude* OR experience* OR barrier* OR facilitator* OR strateg* OR practice* OR behavi*r*) AND (qualitative OR “qualitative research” OR interview* OR “focus group*” OR ethnograph* OR phenomenolog* OR “grounded theory” OR “thematic analysis” OR “content analysis” OR “narrative analysis” OR “discourse analysis”))

We will adapt this search string for each database and document the detailed search strategies in an appendix of the review protocol. The final search strings for each database will be peer-reviewed by a second information specialist using the Peer Review of Electronic Search Strategies (PRESS) checklist [17].

In addition to electronic database searching, we will hand-search the reference lists of all included studies and relevant systematic reviews to identify any additional studies that meet our inclusion criteria. We will also conduct a gray literature search to identify any relevant unpublished studies, theses, or conference proceedings. This will include searching Google Scholar, ProQuest Dissertations & Theses, and websites of relevant organizations such as Health Care Without Harm, Practice Greenhealth, and the Sustainable Development Unit of the UK National Health Service.

### Study selection

Following the search, we will export all identified records to Covidence, a web-based systematic review management platform. Covidence will automatically remove duplicates and allow the review team to collaborate on the study selection process.

Two reviewers will independently screen the titles and abstracts of all unique records against the predefined eligibility criteria. Records that clearly do not meet the inclusion criteria will be excluded at this stage. For records that appear potentially eligible or where eligibility is unclear from the title and abstract, we will obtain the full-text articles. The same two reviewers will then independently assess the full-text articles against the inclusion criteria. Any discrepancies in eligibility decisions between the two reviewers will be resolved through discussion or by consulting a third reviewer if necessary.

We will document the reasons for excluding studies at the full-text stage and present the study selection process using a PRISMA (Preferred Reporting Items for Systematic Reviews and Meta-Analyses) flow diagram [18]. The flow diagram will detail the number of records identified from electronic databases and other sources, the number of duplicates removed, the number of records screened, the number of full-text articles assessed for eligibility, and the final number of studies included in the review, along with reasons for exclusions at each stage.

### Data extraction

Two reviewers will independently extract relevant data from the included studies using a standardized data extraction form within Covidence. Prior to full data extraction, the form will be piloted on a sample of included studies and refined as needed to ensure clarity and consistency. The data extraction form will capture key information including: study characteristics (such as authors, year of publication, country, aim or objective, and setting); participant characteristics (including the type and number of healthcare professionals, age, gender, and years of experience); methodological details (such as study design, sampling strategy, data collection methods, and data analysis approach); study findings (including main themes or categories, supporting quotations, and author interpretations); and conclusions or recommendations relevant to the review question.

Any disagreements in data extraction between the two reviewers will be resolved through discussion or by involving a third reviewer if necessary. If any relevant data are missing or unclear in the included studies, we will attempt to contact the study authors for clarification or additional information.

### Quality appraisal

Two reviewers will independently assess the methodological quality of the included studies using the Critical Appraisal Skills Programme (CASP) checklist for qualitative studies [19]. The CASP checklist consists of 10 questions that address key domains of qualitative research quality, such as the appropriateness of the research design, the rigor of the data collection and analysis methods, the clarity and coherence of the findings, and the relevance and value of the research. Each question is answered as “yes,” “no,” or “can’t tell,” with space for comments to justify the judgments.

The two reviewers will discuss any discrepancies in their quality appraisal assessments and seek to reach consensus. If agreement cannot be reached, a third reviewer will be consulted to arbitrate. We will not exclude studies from the review based on their methodological quality, as this is not recommended for qualitative evidence syntheses [20]. However, we will consider the quality appraisal results when interpreting the findings and drawing conclusions from the synthesis.

### Risk of bias assessment

In addition to the quality appraisal using the CASP checklist, we will assess the risk of bias in the included studies using the Joanna Briggs Institute (JBI) Critical Appraisal Checklist for Qualitative Research [21]. This checklist consists of 10 questions that cover similar domains to the CASP checklist but with a more explicit focus on assessing the potential for bias in qualitative studies.

Two reviewers will independently apply the JBI checklist to each included study, with any disagreements resolved through discussion or by involving a third reviewer. We will present the results of the risk of bias assessment in a tabular format, indicating for each study whether each checklist item was met (yes, no, unclear, or not applicable). As with the quality appraisal, we will not exclude studies based on their risk of bias assessment results but will consider them when interpreting the review findings.

### Data synthesis

We will synthesize the qualitative evidence from the included studies using thematic synthesis, following the approach described by Thomas and Harden [22]. Thematic synthesis is a widely used method for identifying, analyzing, and reporting patterns or themes across qualitative studies, with the aim of generating new insights and interpretations that go beyond the findings of the primary studies.

The thematic synthesis will involve three main stages. First, the two reviewers will independently read and re-read the extracted data from each study, applying line-by-line codes that capture the meaning and content of the text. The reviewers will then compare their initial codes, discuss any discrepancies, and agree on a unified set of codes. In the second stage, the reviewers will work together to group the codes into descriptive themes that closely reflect the main ideas and concepts expressed in the primary study findings. Finally, the reviewers will use the descriptive themes as a basis for generating analytical themes that offer a higher-level interpretation of the data and directly address the review objectives.

Throughout the synthesis process, we will use NVivo 12 qualitative data analysis software to manage the coding and facilitate the organization and comparison of themes across studies. We will present the synthesized findings narratively in the review report, using representative quotations from the primary studies to illustrate key themes and sub-themes. We will also create thematic maps or other visual representations to depict the relationships and connections between the themes.

In conducting the thematic synthesis, we will be mindful of the context and characteristics of each primary study, as well as any methodological limitations or sources of bias identified during the quality appraisal and risk of bias assessment. We will reflect critically on how these factors may influence our interpretation of the findings and the transferability of the synthesized themes to other healthcare settings or populations.

### Confidence in the findings

To assess the confidence that can be placed in the synthesized qualitative findings, we will apply the Grading of Recommendations Assessment, Development and Evaluation Confidence in the Evidence from Reviews of Qualitative research (GRADE-CERQual) approach [23]. GRADE-CERQual provides a systematic and transparent framework for evaluating the extent to which the synthesized findings are a reasonable representation of the phenomenon of interest, based on the methodological limitations, coherence, adequacy, and relevance of the contributing evidence.

For each synthesized finding, we will assess the four components of the CERQual approach to determine our confidence in the evidence. Methodological limitations will be evaluated by examining the extent to which the primary studies contributing to the finding have any quality issues or risk of bias that could reduce our confidence, based on our appraisal results. Coherence will be assessed by considering how consistently the finding appears across the contributing studies and whether any inconsistencies can be explained by contextual or methodological differences. Adequacy of data will involve evaluating the richness and quantity of the data supporting the finding, including the number of studies, depth of information, and whether the data are considered thin or sparse. Relevance will be determined by assessing how closely the contributing studies align with the review question in terms of population, setting, and phenomenon of interest.

For each component, we will make a judgment of “no or very minor concerns,” “minor concerns,” “moderate concerns,” or “serious concerns.” We will then make an overall assessment of the confidence in each synthesized finding as “high,” “moderate,” “low,” or “very low.” The CERQual assessments will be presented in a Summary of Qualitative Findings table, along with an explanation of the judgments made for each finding.

By applying the CERQual approach, we aim to provide a transparent and systematic evaluation of the confidence that can be placed in the synthesized findings, which will help users of the review to interpret the results and assess their applicability to their own contexts.

## Discussion

This systematic review will provide a comprehensive synthesis of qualitative evidence on healthcare professionals’ perceptions and experiences regarding green practices in health services. By exploring the facilitators, barriers, and strategies for implementing sustainable practices from the perspective of healthcare professionals, the review will generate valuable insights that can inform policy, practice, and future research in this important area.

The findings of this review may have several implications for promoting environmental sustainability in healthcare. First, by identifying the factors that enable or hinder the adoption of green practices from the viewpoint of healthcare professionals, the review can help to target interventions and policies to address the most salient barriers and leverage the most effective facilitators. This could include, for example, interventions to improve healthcare professionals’ knowledge and awareness of sustainability issues, policies to incentivize sustainable behaviors, or strategies to engage healthcare professionals in the design and implementation of green initiatives.

Second, by synthesizing healthcare professionals’ experiences with implementing green practices in different healthcare settings, the review can highlight practical challenges and success stories that can inform the development of context-specific sustainability interventions. This may involve adapting existing interventions to fit the needs and constraints of particular healthcare environments or developing new interventions that build on the lessons learned from previous implementation efforts.

Third, by identifying strategies and recommendations for promoting sustainability based on the insights of healthcare professionals, the review can contribute to the development of evidence-based guidelines and best practices for green healthcare. Such guidelines could help healthcare organizations to prioritize and operationalize sustainability initiatives, as well as provide a benchmark for evaluating the effectiveness of these initiatives over time.

More broadly, this review will contribute to advancing the theoretical and conceptual understanding of sustainability in healthcare. By synthesizing qualitative evidence from a range of healthcare contexts and professional perspectives, the review may identify common themes, patterns, or mechanisms that underlie the adoption and implementation of green practices in healthcare. These insights could inform the development of new theoretical frameworks or models that explain how and why sustainability interventions work (or fail to work) in healthcare settings, and how they can be optimized to achieve desired outcomes.

However, it is important to acknowledge the potential limitations of this review. First, by including only studies published in English, we may miss relevant evidence from other linguistic and cultural contexts, which could limit the generalizability of the findings. Second, the qualitative nature of the evidence may not allow for definitive conclusions about the effectiveness or impact of specific green practices, as qualitative studies are not designed to quantify outcomes or establish causal relationships. Third, the synthesis will depend on the depth and quality of the primary studies, which may vary in terms of their methodological rigor and reporting clarity.

Despite these limitations, we believe that this systematic review will make a valuable contribution to the growing field of sustainable healthcare. By focusing on the perspectives and experiences of healthcare professionals, the review will complement existing research on the technical, operational, and environmental aspects of green practices in healthcare. It will also respond to calls for more interdisciplinary and stakeholder-engaged approaches to sustainability research, recognizing that the successful implementation of green practices depends not only on technological solutions but also on the buy-in and participation of healthcare professionals and other key stakeholders.

The findings of this review will be disseminated through multiple channels to maximize their reach and impact. First, we will publish the completed review in a peer-reviewed academic journal focused on sustainability, environmental health, or healthcare management. Second, we will present the results at relevant national and international conferences, such as the CleanMed conference or the International Conference on Environmental, Cultural, Economic, and Social Sustainability. Third, we will prepare a policy brief summarizing the key findings and recommendations for distribution to healthcare organizations, professional associations, and policymakers. Finally, we will engage with local healthcare providers and sustainability champions to discuss the implications of the findings for their own organizations and to identify opportunities for collaborative research or implementation projects.

In conclusion, this systematic review of qualitative evidence on healthcare professionals’ perceptions and experiences regarding green practices in health services will provide valuable insights into the factors that influence the adoption and implementation of sustainable practices in healthcare. By synthesizing evidence from the perspective of key stakeholders, the review will identify strategies for overcoming barriers, leveraging facilitators, and engaging healthcare professionals in the transition to more environmentally sustainable models of care. Ultimately, we hope that this review will contribute to the development of evidence-based policies and practices that can help to reduce the environmental impact of healthcare while maintaining or improving the quality and accessibility of care for patients and communities.

## Data Availability

All data produced in the present study are available upon reasonable request to the corresponding author

## Appendix 1 Data extraction form

Appendix 1: Data extraction form
| Category | Items |
| --- | --- |
| **1. Study Characteristics** | Authors<br>Year of publication<br>Country<br>Aim/objective<br>Setting (e.g., hospital, primary care, community) |
| **2. Participant Characteristics** | Type of healthcare professionals (e.g., doctors, nurses, managers)<br>Number of participants<br>Age (mean, range)<br>Gender (% female)<br>Years of experience (mean, range) |
| **3. Methodology** | Study design (e.g., interviews, focus groups, ethnography)<br>Sampling strategy (e.g., purposive, snowball, convenience)<br>Data collection methods (e.g., semi-structured interviews, observation)<br>Data analysis approach (e.g., thematic analysis, grounded theory) |
| **4. Findings** | Main themes or categories reported<br>Supporting quotations<br>Author interpretations |
| **5. Conclusions and Recommendations** | Key conclusions relevant to the review question<br>Recommendations for policy, practice, or research |
| **6. Miscellaneous** | Funding source<br>Conflicts of interest<br>Limitations acknowledged by authors |

